# A study protocol for community use of digital auscultation to improve diagnosis of paediatric pneumonia in Bangladesh

**DOI:** 10.1101/2021.11.18.21266302

**Authors:** Salahuddin Ahmed, Dipak Kumar Mitra, Harish Nair, Steve Cunningham, Ahad Mahmud Khan, Md. Ashraful Islam, Ian McLane, Nabidul Haque Chowdhury, Nazma Begum, Mohammod Shahidullah, Sariful Islam, John Norrie, Harry Campbell, Aziz Sheikh, Abdullah H. Baqui, Eric D. McCollum

## Abstract

**Introduction:** The World Health Organisation’s Integrated Management of Childhood Illnesses (IMCI) algorithm relies on counting respiratory rate and observing respiratory distress to diagnose childhood pneumonia. IMCI performs with high sensitivity but low specificity, leading to over-diagnosis of child pneumonia and unnecessary antibiotic use. Including lung auscultation in IMCI could improve pneumonia diagnosis. Our objectives are: (i) assess lung sound recording quality by primary health care workers (HCWs) from under-five children with the Feelix Smart Stethoscope; and (ii) determine the reliability and performance of recorded lung sound interpretations by an automated algorithm compared to reference paediatrician interpretations.

**Methods and analysis:** In a cross-sectional design, Community HCWs will record lung sounds of ∼1,000 under-five-year-old children with suspected pneumonia at first-level facilities in Zakiganj sub-district, Sylhet, Bangladesh. Enrolled children will be evaluated for pneumonia, including oxygen saturation, and have their lung sounds recorded by the Feelix Smart stethoscope at four sequential chest locations: two back and two front positions. A novel sound-filtering algorithm will be applied to recordings to address ambient noise and optimize recording quality. Recorded sounds will be assessed against a pre-defined quality threshold. A trained paediatric listening panel will classify recordings into one of the following categories: normal, crackle, wheeze, crackle and wheeze, or uninterpretable. All sound files will be classified into the same categories by the automated algorithm and compared with panel classifications.

**Conclusions:** Lung auscultation and reliable interpretation of lung sounds of children are usually not feasible in first-level facilities in Bangladesh and other low- and middle-income countries (LMICs). Incorporating automated lung sound classification within the current IMCI pneumonia diagnostic algorithm may improve childhood pneumonia diagnostic accuracy at LMIC first-level facilities.

**Ethics and dissemination:** Ethical review has been obtained in Bangladesh (BMRC Registration Number: 09630012018) and in Edinburgh, Scotland, United Kingdom (REC Reference: 18-HV-051). Dissemination will be through conference presentations, peer-reviewed journals and stakeholder engagement meetings in Bangladesh.

**Trial registration number:** NCT03959956

**Article summay:** Strengths and limitations of this study

- Evaluating the quality of lung sound recordings in a first-level facility where auscultation is usually unavailable and challenging to obtain due to a typically crowded and noisy environment and providers may not get enough time to calm the child due to time pressure from a high-volume patient.
- This study will assess the feasibility of recording lung sounds by front line community health workers who do not usually use conventional stethoscopes during clinical care.
- Two standardised paediatricians masked to the child’s clinical status will independently classify the recorded lung sounds, and a third masked and independent paediatrician will arbitrate any discrepancies.
- A machine-learning algorithm developed by Johns Hopkins and Sonavi Labs will detect abnormal lung sounds and be compared with classifications by human listeners/paediatricians.
- The study will not have chest radiography findings of enrolled children, which is considered by many a gold standard for pneumonia diagnosis, as chest radiography is not available at this level of the health system in Bangladesh. Instead, this study will measure the peripheral oxyhaemoglobin saturation and evaluate clinical examination findings, including respiratory danger signs data.

## INTRODUCTION

Childhood pneumonia is one of the leading causes of death in children younger than five years globally^1^ and accounts for an estimated 0.8 million deaths in children annually^2^. The World Health Organization (WHO) estimates that the African and the South-East Asian Regions contribute to more than 75% of total paediatric deaths from pneumonia^3^. It is estimated that the annual incidence of childhood pneumonia in low- and middle-income countries (LMICs) is 231 episodes per 1,000 children^3^. Pneumonia is also a significant cause of hospitalisation^4^, and about 16.4 million children in LMICs were hospitalised due to pneumonia in 2015^3^. A population-based study in Bangladesh estimated that the annual incidence of pneumonia was 360 episodes per 1,000 child-years, of which 7.3% were hospitalised^5^. The case-fatality rate of child pneumonia in Bangladesh is estimated to be 2-4% ^6 7^.

The World Health Organization (WHO) and United Nations Children’s Fund (UNICEF) Integrated Management of Childhood Illness (IMCI) guidelines have been the foundation of pneumonia management in LMICs since mid-90s ^8 9^. Per current IMCI guidelines, a child with fast breathing and/or chest indrawing without any danger sign is classified as non-severe pneumonia, and treated at home with oral antibiotics^10^. These guidelines have proven to be one of the most important childhood pneumonia interventions for LMICs to date, and up to 36% of the under-five-year-old mortality reductions in LMICs have been attributed to guideline implementation^11-15^.

Despite its overall success, the IMCI algorithm can still be improved. Firstly, the guidelines were developed when access to vaccines was limited and child pneumonia mortality was high, so the guidelines intentionally prioritised sensitivity over specificity to ensure children with possible bacterial disease received antibiotics^8 10^. Thus, the IMCI algorithm over-diagnoses many children with pneumonia who do not require antibiotics and they may have different treatable diseases^16 17^. Secondly, the WHO guidelines do not include lung auscultation in their pneumonia definition for frontline healthcare workers despite lung auscultation serving as the cornerstone for pneumonia diagnosis in most ambulatory, well-resourced settings staffed by clinicians trained to perform lung auscultation ^10^. The exclusion of auscultation findings in these guidelines likely stems from its high inter-observer variability and subjectivity, regardless of the training level of healthcare providers, and the related difficulty in training healthcare workers to effectively use a conventional stethoscope^18-22^. Traditional stethoscopes themselves are also limiting, attenuating higher frequency sounds, like wheezing and crackles, yet transmitting ambient noises and tubular resonance effects^20 23 24^. Automated real-time classification of lung sounds or digital auscultation may overcome these limitations^25^. Digital auscultation augmented by artificial intelligence (AI) algorithms has the potential to be a highly specific respiratory diagnostic tool feasible for use by first-level healthcare workers in LMICs. Operationally, the inclusion of adventitious sound classifications in current IMCI guidelines could help to reduce ^26^ unnecessary use of antibiotics in LMICs.

A digital stethoscope can convert an acoustic sound to electronic signals, which can be further amplified for optimal listening. These electronic signals can then be processed and digitalised to transmit to a personal computer or a laptop^27^. Automatic lung sound analysis, aiming to overcome the limitations of conventional auscultation, has been the recent focus of a significant amount of research, and some commercial systems are already available in the market^28 29^.

The digital stethoscope has advantages over the analogue stethoscope in different stages of auscultation. An analogue stethoscope requires the proper placing of the diaphragm or bell in the correct positions of the human body to listen to internal body sounds. More modern digital stethoscopes do not necessarily require exact placement for two reasons. First, they convert the acoustic wave into electric signals and replace the double-sided chest piece (diaphragm and bell) with transducers to convert acoustic signals to electric signals. Second, the chest piece is packed with transducer arrays to achieve a uniform sensitivity over the entire active area. Together this design delivers a strong signal even when the chest piece is not placed in precisely the right position^30 31^. This advantage is essential for minimally trained health care providers.

Conventional auscultation using an analogue stethoscope also requires a quiet environment, and ideally with the patient in a quiet, cooperative state, which is difficult in hospitals/clinics and especially hospitals/clinics in LMICs where the number of patients is typically usually higher than capacity. This often results in patient examination rooms filled with chattering people, ringing phones, and whirring fans; most importantly, the child being examined may then be agitated, uncooperative, and / or crying. Digital stethoscopes could improve listening capability through the use of noise cancelling technologies^32 33^. Limitations of the human auditory system, which ranges from 20 Hz to 20 kHz for young adults and the range shrinks after middle-age^34^, is also a drawback in conventional auscultation using a conventional stethoscope. A digital stethoscope can amplify sounds up to 100 fold^35^. Much experience is required to interpret body sounds, and inter-rater variability is high regardless of healthcare providers’ training level^21^. Machine learning techniques can be used in digital auscultation to auto-analyse sounds^28^ and produce a diagnosis or treatment decision^23^. Johns Hopkins University and Sonavi Labs developed a novel digital stethoscope named Feelix Smart Stethoscope (Figure: 1)^30 31^, which improves lung signal strength by uniformly distributing highly sensitive microphones in an array pattern across the stethoscope diaphragm to increase the sensitivity and provide broader frequency response, a critical feature for identifying higher frequency pathologic lung sounds. Its 3.7 V and 250 amps/hour rechargeable battery can power >20 hours of use, important in rural communities with unreliable electricity. The device mitigates movement artefact and tubular resonance by using an ergonomic design to better secure the device on the child’s chest. It also eliminates the rubber stethoscope tubing, a source of ambient noise and friction contamination. Notably, the device includes an integrated external-facing microphone that captures simultaneous environmental noise to the lung sound recording and removes the unwanted ambient noises through an adaptive spectral subtraction schema^24 36^. The Feelix Smart Stethoscope also permits onboard data storage with a microSD card. The device turns on when the user picks up the device or touches the top of the device. The device turns off automatically after 60 seconds when it does not sense any touch. The device has three touch buttons – button-1 to start recording, button-2 to start a new session to record a new child’s four chest points recording, and button-3 to establish a Bluetooth connection with a mobile phone or tablet. It also has a slider to control sound volume level. In each session, the device automatically records 10 seconds for each four chest points of a child. The Feelix Smart Stethoscope has been successfully validated in the laboratory against six other commercially available electronic stethoscopes – including the Littmann 3200 electronic stethoscope and Thinklabs – and has demonstrated comparable results^37 38^. Johns Hopkins and Sonavi Labs developed a machine-learning algorithm that can provide an automated classification of adventitious lung sounds. This Feelix Smart Stethoscope and the machine-learning algorithm will be used in this study.

**Figure 1:**
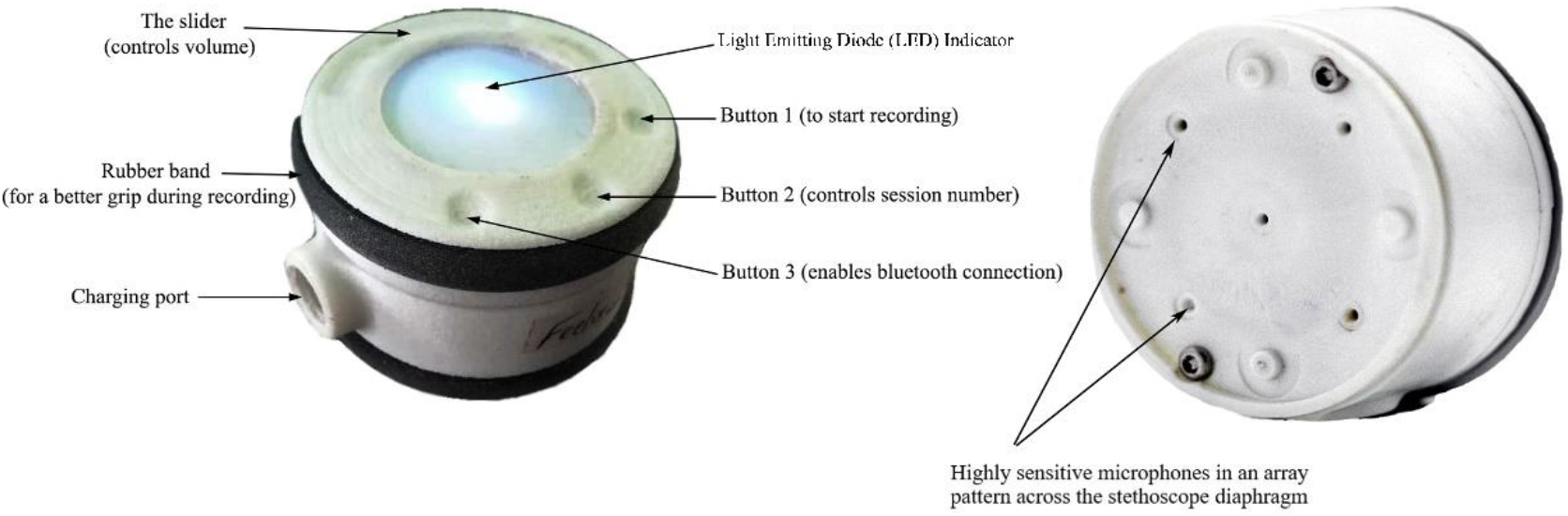
Feelix Smart Stethoscope

### Objectives and hypotheses

#### The primary objectives of the study are

i. to assess whether lung sounds recorded using the Feelix Smart Stethoscope in children by health workers at community level health facilities meet pre- defined quality thresholds established by experts; and
ii. to determine the reliability and performance of the Smartscope Respiratory Detector automated analysis algorithm on lung sounds recorded by community health care provider (CHCP) using the Smartscope, compared to reference interpretations by a paediatric listening panel.

#### The study hypotheses are

i. More than 50% of patients will have ‘quality’ lung sound recordings (targeted goal), defined as at least 75% interpretable lung sound segments per patient (i.e., 3 out of 4 chest positions).
ii. (ii) The agreement between automated computerised analysis (respiratory detector) and paediatric listening panel will be high (kappa >0.8).

## METHODS AND ANALYSIS

### Study setting

This study will be implemented in the Projahnmo field site, a site for maternal, new-born, and child health research, which was established in 2001 in Sylhet district of Bangladesh by a partnership of Johns Hopkins University, the Bangladesh Ministry of Health and Family Welfare (MOHFW), and several Bangladeshi institutions, including Non-Government Organisations (NGOs) and academia. A well-established routine community-based pregnancy, birth and under-five child surveillance system are being maintained by trained female community health workers (CHWs) in this site. The CHWs identify sick children during routine household visits and refer the children to the health facilities. They also educate carers of children about pneumonia signs and symptoms so that the carers can visit nearby community clinic (CC) without delay.

Bangladesh has established about 13,000 CCs, one each for ∼6,000 people^39^. Each CC is staffed by a CHCP with at least 12^th^ grade education and three months of pre-service training, including IMCI guidelines. Each CHCP is responsible for providing primary health care for the population, including its catchment area’s children. Nine CCs will be purposively selected from the Projahnmo surveillance area (Zakiganj sub-district of Sylhet district of Bangladesh) (Figure: 2). CHCP of respective CC will screen all under-5 children while providing primary health care.

**Figure 2:**
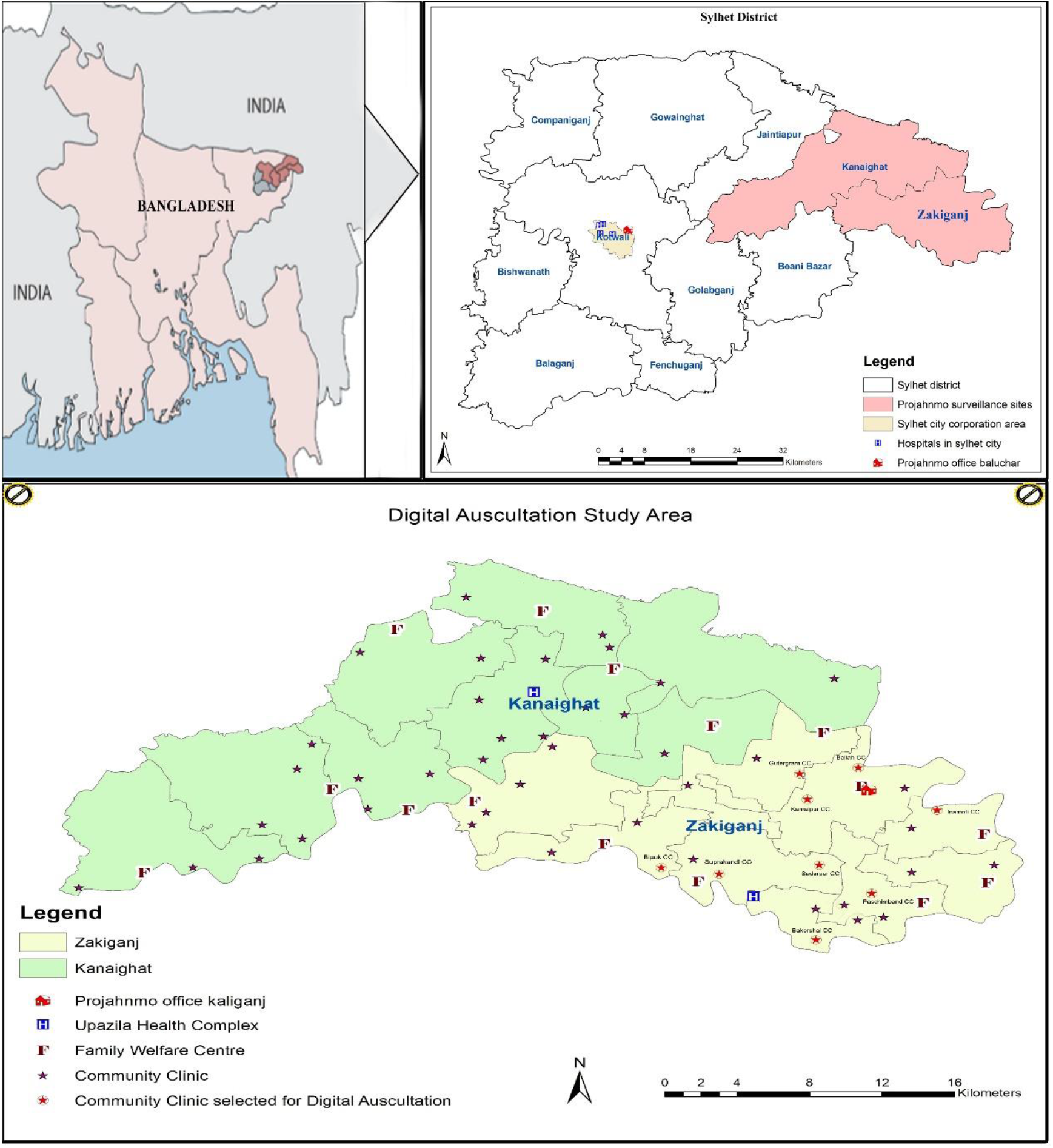
Study site

All CHW and CHCP will be trained and standardised to identify signs and symptoms of pneumonia according to WHO IMCI guidelines. A study physician will be recruited for providing training and supervision of CHCP and CHW in clinical assessments, measurement of peripheral oxyhaemoglobin saturation (SpO_2_), and recording of lung sounds using the Smartscope. However, the study physician will not be directly involved in recording lung sound from the study participants.

### Study design and procedure

Using a cross-sectional design, each CHCP will screen all children younger than five years visiting the community clinic. CHCP will obtain consent and record lung sounds of children who fulfilled the following criteria: history and/or observed cough, and/or history and/or observed difficult breathing, and a permanent resident of the Projahnmo site and did not enrol in the study within the past 30 days. CHCP will record sounds from four chest locations – two from the back and two from the front (Figure: 3). Each position will be recorded for approximately 10 seconds and the overall recording process will take about one minute. The recorded sound files will be then transferred to a password-protected server. The CHCP will also examine the child f or fast breathing by manually counting respirations and observe for abnormal breathing patterns such as lower chest wall indrawing, nasal flaring, head nodding, tracheal tugging, grunting, intercostal retractions, and stridor when calm. CHCP will measure the SpO_2_ using a Masimo® Rad5 pulse oximeter, temperature using a digital thermometer, and anthropometry (weight, height and mid-arm circumference) using standard tools and techniques. The SpO_2_ data will be used to classify pneumonia according to IMCI guidelines. If any child has a SpO_2_ <90%, then referral to the sub-district health centre or Sylhet Osmani Medical College Hospital will be initiated. All enrolled children will be assessed after day-8 of enrolment by Projahnmo CHWs for treatment compliance and treatment outcome.

**Figure 3:**
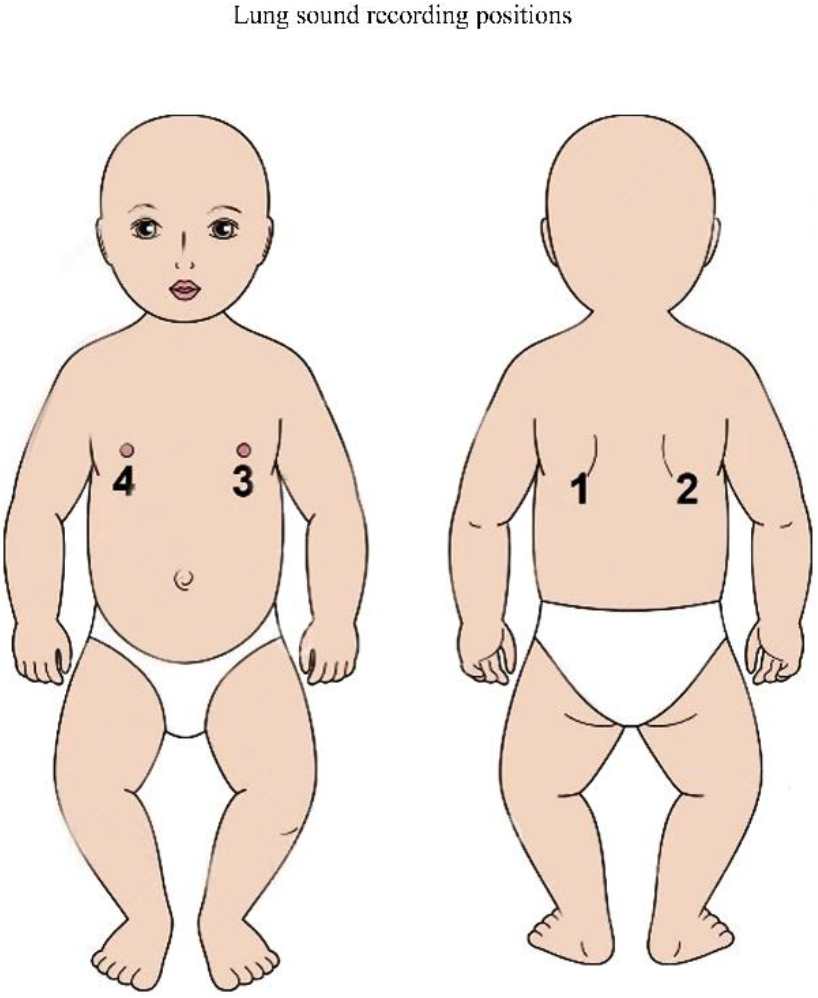
Sound recording positions

A total of 11 paediatricians will be trained to the methodology developed and validated during the Pneumonia Etiology Research for Child Health (PERCH) study^40^. Only paediatricians successfully standardised to the methodology will serve as human listening panel members. Panellists will classify the recorded sound files of all four positions separately into five categories, e.g. (1) no wheeze and no crackles, (2) wheeze only (no crackles), (3) crackles only (no wheeze), (4) both wheeze and crackles or (5) uninterpretable. Two primary listeners will independently classify the recorded lung sounds, and any discrepancies will be arbitrated by the third listener (EDM). Johns Hopkins and Sonavi Labs developed a machine-learning algorithm; all sound files will also be classified the same categories using this algorithm and will be compared with human classification. Figure 4 depicts the study flow.

**Figure 4:**
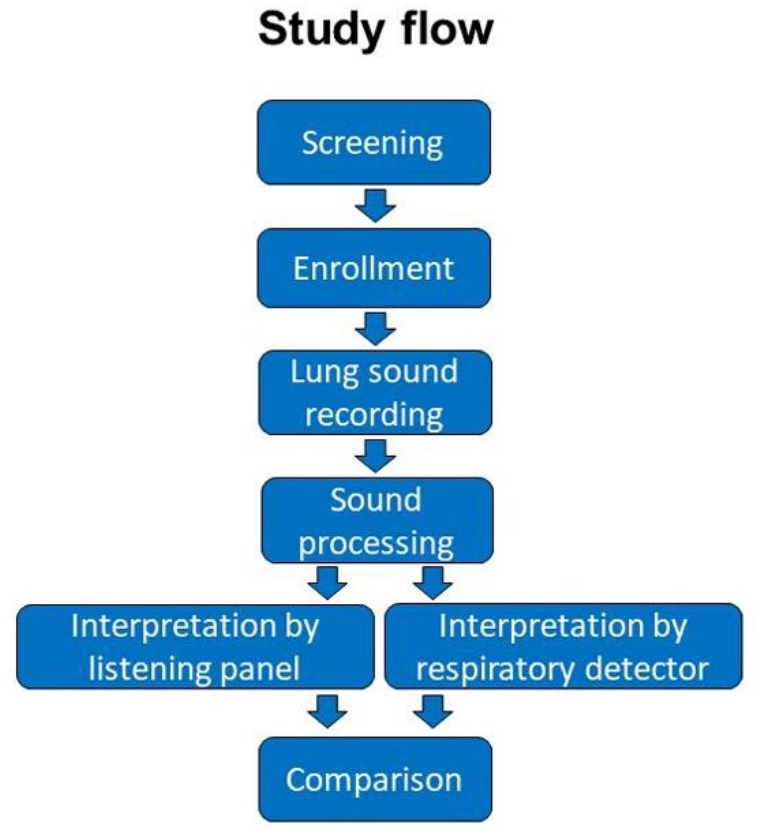
Study flow

### Sample size calculation

Cohen’s kappa will be used to assess inter-listener and computer-human agreement in this study. To detect a true kappa of 0.50 compared to a kappa of 0.40 under null hypothesis and assuming two-sided type I error of 5%, power of 80% and two categories of frequencies equal to 30% and 70% normal and abnormal findings respectively, a sample size of 752 will be required.^41^ This sample size has been calculated in Power Analysis and Sample Size (PASS) software version 11.

We will include all quality recordings for this assessment. A quality recording is defined as a recording with >75% interpretable chest positions (i.e., 3 of the 4 chest positions recorded are interpretable). Assuming that 75% of all recordings will be of acceptable quality, we will need 1,003 subjects to be enrolled in this study to obtain our required sample size of 752.

### Statistical analysis

A point estimate for the proportion of recordings along with a 95% confidence interval that meets the definition of quality will be determined. The numerator will be the total number of recordings that were interpreted to meet quality criteria and the denominator will be the total number of recordings collected and interpreted. A raw agreement percentage will be determined between the interpretations of the two primary human listeners of the listening panel. The numerator will be the total number of recordings with an interpretation from the first and second human listener that agree, and the denominator will be the total number of recordings that have a final human interpretation from both primary listeners. Cohen’s kappa, a metric that assesses the agreement beyond chance will be calculated, and a kappa adjusted for prevalence and bias will also be calculated. A kappa above 0.8 will be defined as perfect agreement, 0.61-0.8 as high agreement, 0.41-0.6 as moderate agreement, 0.21-0.4 as fair agreement, 0.01-0.2 as low agreement, and 0 as no agreement.

A raw agreement percentage between the overall human listening panel’s final interpretation result and the computerised analysis algorithm’s final interpretation result will be determined. The numerator will be the total number of recordings with an interpretation from the human listening panel and the computerised algorithm that agree, and the denominator will be the total number of recordings that have both a human and computerised interpretation. Cohen’s kappa, a metric that assesses the agreement beyond chance will be calculated, and a kappa adjusted for prevalence and bias will also be calculated. The same kappa scale detailed previously will be used for interpretation.

The performance of the computerised interpretation algorithm will be evaluated when assuming the human listening panel’s final interpretation is the gold standard. Sensitivity, specificity, positive predictive value, negative predictive value, and positive and negative likelihood ratios will be assessed.

### Patient and public involvement

We formed Patient Public Involvement Groups (PPIG) consisting of CHCPs, CHWs, Health Assistants, Physicians, community leaders, religious leaders, parents of under-5 children, teachers, local journalists and organised several meetings in the community for their insights, approval, and support to implement the study. They believe its feasible and acceptable to implement the study. We also consulted and engaged several district and national level stakeholders including public health program managers, policymakers, paediatricians, physicians, and civil society representatives during the development of the protocol and implementation strategy of the study. A technical review committee (TRC) consisting of policymakers and technical experts formed by the Bangladesh MOHFW reviewed and approved the protocol.

The study results will be reviewed by the PPIG and TRC before publishing in peer-reviewed journals, presented at international conferences and to health officials in Bangladesh. The findings also will be disseminated to stakeholders at Zakiganj sub-district, Sylhet district and national level at Dhaka, Bangladesh.

### Ethics and registration

Ethical approval is obtained from the National Research Ethics Committee of Bangladesh Medical Research Council (BMRC), Bangladesh (Registration Number: 09630012018), and Academic and Clinical Central Office for Research and Development (ACCORD) Medical Research Ethics Committee (AMREC) NHS, Lothian, Edinburgh, UK (REC Reference: 18-HV-051). This study is registered with ClinicalTrails.gov (NCT03959956).

### Data collection, storage, security and sharing

Data will be collected using password-protected electronic devices (Samsung Galaxy Tab A 7.0) in the Android platform. The data will be transferred to a server (SQL Server 2008 R2) located at Sylhet, Bangladesh in real-time using internet connectivity and will keep a backup copy daily in another server at the study Dhaka office. All the tablets and servers will be password protected. Recorded lung sounds will be transferred to the server. Paediatric listening panel members will fill up the lung sound interpretation on an online database which will also be stored on the server in real-time. Access to collected data will be restricted to individuals from the research team treating the participants, representatives of the sponsor(s), and representatives of regulatory authorities. Anonymised data files will also be stored securely in the DataStore repository at the University of Edinburgh, UK and will be shared after publication of main paper.

## DISCUSSION

This study aims to demonstrate the feasibility of collecting quality lung sounds by frontline health workers and to examine the performance of the machine learning algorithm against a panel of human listeners for identifying adventitial lung sounds. To our knowledge this is the first study that will assess the recording of lung sounds of children younger than five years in first-level facilities by frontline workers in LMICs, where the burden of pneumonia and antibiotic use is high and diagnostic capacity is limited. The PERCH study enrolled children at a hospital setting and digital auscultation was performed by physicians, formally trained clinical assistants, or nurses ^40^, another study enrolled children at a tertiary level centre in Lima, Peru^42^, and paediatricians recorded lung sounds in two tertiary level teaching hospitals in a study in Nepal^43^. Recording lung sounds in a first-level facility poses unique challenges in that clinics are typically crowded and the environment can be chaotic and noisy. Furthermore, ill children younger than five years of age can be especially uncooperative in uncomfortable ambulatory settings, leading to unique challenges like agitation, crying, vocalizations, and the associated auscultation artifacts these create. Healthcare providers in ambulatory settings may lack the necessary time to calm the child and address these issues due to pressures from a high patient volume. If frontline health workers can effectively use digital auscultation in their typical clinical setting, then many false-positive pneumonia cases may be spared from treatment with antibiotics, which may reduce the cost of treatment as well as reduce the chance of developing antimicrobial resistance. A systematic analysis of 132 national surveys from 73 countries reported that on average, four of ten ill children below age five years in LMICs are treated with antibiotics^44^. Recently, United Nations General Assembly announced that antimicrobial resistance is the most important and urgent threat globally^45^.

One limitation of our study is that chest radiography will not be performed as it is not routinely available in first-level facilities in Bangladesh, and we will not compare radiographic imaging with lung sound classification. Instead, lung sound classifications will be compared with clinical findings among children meeting IMCI pneumonia to those who do not. Lack of a gold standard reference for pneumonia is well understood, and it is widely accepted that chest radiography itself lacks diagnostic accuracy for pneumonia^46^. For example, it was found that many children with IMCI defined clinical pneumonia (age-specific tachypnoea) had normal chest radiographs^47^ and may have normal lung sounds and may not require antibiotics. In this study we will utilize the paediatrician listening panel as our gold standard reference as previously described^40^. We have shown that lung sound classifications of digital auscultation recordings generated using a listening panel approach has strong associations with radiographic findings and also mortality outcomes^48^.

Future work ranges from additionally refining this device, based on lessons learned from the use of the device at the first level facility during this study, as well as phase 1 and 2 clinical trials that prospectively integrate the digital auscultation into the WHO pneumonia management pathway to evaluate patient outcomes after digital auscultation-based decision-making. Eventually, if these preliminary studies are successful, a large multi-country clinical trial could be designed to evaluate the safety and efficacy of digital auscultation to improve pneumonia management.

## Data Availability

Anonymised data files will also be stored securely in the DataStore repository at the University of Edinburgh, UK and will be shared after the publication of the main paper.

## Authors contributions

EDM, SA and AHB conceptualised and designed this study. AHB, EDM, HN, SC provided mentorship to SA. HC, AS and DKM critically review the study design. JN and DKM reviewed the sample size estimation and analysis plan. EDM will provide training to the human listening panel and IM will analyse the recorded sound files using a machine learning algorithm. NHC and NB will be responsible for data management and AI will manage the field implementation of this study. SA drafted the manuscripts and all authors critically reviewed and approved the final manuscript before submission.

## Acknowledgements

The authors are grateful for the support and contributions of Dr Arunangshu Dutta Roy, Dr Arifa Islam, Dr Iffat Ara Jaben, Md. Shafiqul Islam, Asim Nehal, Dr. Md. Shamsul Haque, Dr. Himangshu Lal Roy, Dr. Premananda Mondol, Dr. Md. Jahurul Islam, Dr. Sabina Ashrafee Lipi, Dr. Abdullah Al Mehedi, Dr. Husam Md. Shah Alam, and the Ministry of Health and Family Welfare, Government of Bangladesh.

